# Cortical re-organization after traumatic brain injury elicited using functional electrical stimulation therapy: A case report

**DOI:** 10.1101/2020.05.31.20118323

**Authors:** Matija Milosevic, Tomoya Nakanishi, Atsushi Sasaki, Akiko Yamaguchi, Taishin Nomura, Milos R. Popovic, Kimitaka Nakazawa

## Abstract

Functional electrical stimulation therapy (FEST) can improve motor function after neurological injuries. However, little is known about cortical changes after FEST and weather it can improve motor function after traumatic brain injury (TBI). Our study examined cortical changes and motor improvements in one male participant with chronic TBI suffering from mild motor impairment affecting the right upper-limb during 3-months of FEST and during 3-months follow-up. In total, 36 sessions of FEST were applied to enable upper-limb grasping and reaching movements. Short-term assessments carried out using transcranial magnetic stimulation (TMS) showed reduced cortical silent period (CSP), indicating cortical and/or subcortical inhibition after each intervention. At the same time, no changes in motor evoked potentials (MEPs) were observed. Long-term assessments showed increased MEP corticospinal excitability after 12-weeks of FEST, which seemed to remain during both follow-ups, while no changes in CSP were observed. Similarly, long-term assessments using TMS mapping showed larger hand MEP area in the primary motor cortex (M1) after 12-weeks of FEST as well as during both follow-ups. Corroborating TMS results, functional magnetic resonance imaging (fMRI) data showed M1 activations increased during hand grip and finger pinch tasks after 12-weeks of FEST, while gradual reduction of activity compared to after the intervention was seen during follow-ups. Widespread changes were seen not only in the M1, but also sensory, parietal rostroventral, supplementary motor, and premotor areas in both contralateral and ipsilateral hemispheres, especially during the finger pinch task. Drawing test performance showed improvements after the intervention and during follow-ups. Our findings suggest that task-specific and repetitive FEST can effectively increase cortical activations by integrating voluntary motor commands and sensorimotor network through FES. Overall, our results demonstrated cortical re-organization and improved fine motor function in an individual with chronic TBI after FEST.

## 1. Introduction

Acquired brain injuries, such as stroke or traumatic brain injury (TBI), can cause large portions of the frontal and parietal cortex and/or subcortical structures such as the striatum and thalamus to be affected, which can induce sensorimotor impairment in the contralateral limbs (Nudo et al. 2013). Neurological injuries resulting from trauma are typically diffuse and affect widespread cortical activation changes associated with movement of the paretic limbs. Even in case of focal brain injuries, disruption of sensorimotor networks can trigger reassembly of inter- and intra-cortical networks, resulting in loss of fine motor control (Nudo et al. 2013). Excitability of the motor cortex can be considerably reduced near the injury site, resulting in decreased cortical motor map representations of the affected muscles (Traversa et al. 1997; Butefisch et al. 2006). Spontaneous (natural) recovery can occur even in absence of rehabilitative intervention in the acute stages (Nudo et al. 2013). Compensating behaviours and learned non-use can also arise if unsuccessful attempts to use affected limbs persist (Taub et al. 1998). By restraining use of the non-affected limb, constraint-induced movement therapy has been shown to improve use of the affected limb (Wolf et al. 2006). Intact motor areas adjacent to the injury site and areas outside of the motor cortex or ipsilateral cortical areas may contribute to recovery via intracortical connectivity networks (Weiller et al. 1992; Seitz et al. 2005; Nudo et al. 2013). However, enabling successful movement execution of the affected limbs is still challenging.

Functional electrical stimulation (FES) is a neurorehabilitation approach that can be used to apply short electric impulses on the muscles to generate muscle contractions in otherwise impaired limbs with the goal of assisting motor function (Popovic et al. 2002; Quandt and Hummel 2014; Carson and Buick 2019). When stimulation is sequenced over the appropriate muscles, FES can generate functional movements, including grasping and reaching (Popovic et al. 2001; Popovic et al. 2002). Applications of FES include improving voluntary limb movements in individuals such as stroke and incomplete spinal cord injury (SCI). Specifically, using FES therapy or FEST (Popovic et al. 2002), we have previously demonstrated recovery of upper-limb function in a randomized control trial with stroke patients (Thrasher et al. 2008). FEST was delivered along with conventional therapy in the intervention group, while the control group received 45 min of conventional therapy for 3 to 5 days per week for a total of 12 to 16 weeks (40 sessions in total). Compared to the control group, the stroke FEST group improved in terms of object manipulation, palmar grip torque, and pinch grip force (Thrasher et al. 2008). Another randomized trial with cervical incomplete SCI individuals tested short- and long-term efficacy of 60 min of FEST applied for 5 days per week for 8 weeks (40 sessions), over conventional occupational therapy for improving voluntary upper-limb function (Kapadia et al. 2011). Participants receiving FEST showed greater improvements in hand function at discharge, as well as at 6-month follow-up, compared to the control group (Kapadia et al. 2011). Therefore, FEST was shown as an effective treatment to improve voluntary upper-limb motor function in individuals with both acute and chronic neurological injuries. Despite the clinical evidence, little is known about cortical changes after FEST and whether it can be effective for treating motor dysfunction after TBI.

Repetition, temporal coincidence, and context-specific reinforcement during motor task performance can help induce experience-dependant cortical plasticity after TBI (Nudo et al. 2013). During FEST, task-specific and repeated training is delivered with the assistance of a therapist. Specifically, participants are first asked to attempt to perform a motor task, while the therapist provides reinforcement by triggering appropriate muscles using FES to assist completion of attempted tasks (Popovic et al. 2002). FEST can therefore deliver sensorimotor integration-based training which can help guide experience-dependant cortical plasticity after TBI. Nonetheless, reports on FEST after TBI are relatively few and far between. While some studies showed possible effectiveness of FES for motor recovery after TBI (Oostra et al. 1997; McCain and Shearin 2017), conflicting results have also been shown in a randomized trial (de Sousa et al. 2016). Therefore, the objective of the current study was to investigate the efficacy of the FEST using protocols developed by our team (Thrasher et al. 2008; Kapadia et al. 2011) on improving upper-limb motor function and cortical re-organization in a clinical case study with an individual suffering from mild upper-limb motor impairment after chronic TBI. Specifically, the objectives were to understand cortical changes using neuroimaging and neurophysiological evaluations as well as to examine motor function changes during FEST. Based on our results in stroke (Thrasher et al. 2008) and incomplete SCI (Kapadia et al. 2011), we hypothesized that FEST would be effective to improve upper-limb motor function, which would be accompanied by cortical changes after the therapy.

## 2. Materials and Methods

### 2.1. Clinical presentation

A participant was a male in his late 30’s with a TBI resulting from a motor vehicle accident. The accident occurred 7 years prior to start of the study. At the onset of the study, the participant was diagnosed by his medical team with symptoms of mild motor impairment affecting the right upper- and lower-limbs and higher brain dysfunction, which were the results of the TBI (see Supplementary information: Participant history). The participant was enrolled in the study aiming to improve upper-limb function using FEST. The participant was informed about the study objectives and signed a written informed consent in accordance with the principles of the Declaration of Helsinki. The study protocol was approved by the local institutional research ethics committee at the University of Tokyo.

### 2.2. Functional electrical stimulation therapy

FES was delivered using the Complex Motion system (Compex, Switzerland). Electrical stimulation was used to activate the muscles by applying a rectangular, biphasic, asymmetric charge balanced stimulation pulses at a frequency of 40 Hz and 300 µsec pulse width (Popovic et al. 2001; Popovic et al. 2002). Electrical stimulation was applied on the muscles using surface electrodes (5×5 cm square electrodes on larger muscles and 2 cm diameter circular electrodes on smaller muscles). During each FEST session, the therapist determined the stimulation levels for each muscle by gradually increasing the FES amplitude in 1 mA increments until they identified palpable contractions. The stimulation amplitude was then set to 150% of the amplitude that evoked palpable contractions, and adjusted if necessary, to produce smooth muscle contractions (for average amplitudes, see Supplementary information: FES).

The FEST training protocol is summarized in Figure 1. Training was delivered over the course of 3-months (12-weeks), with 3 sessions per week, each lasting 45 to 60 min (Figure 1A). Each FEST session consisted of three functional training protocols, consistent to previous FEST protocols (Thrasher et al. 2008 and Kapadia et al. 2011), which are illustrated in Figure 1B (see Supplementary information: FES). In each protocol, participant performed a specific functional task, including grasping a water bottle (palmar grasp), bringing an object to his mouth (hand-mouth), and pointing towards a target (pointing forward). For each trial, the participant was first asked to attempt to perform the task, while the therapist triggered a pre-programmed FES sequence to assist voluntary efforts.

**Figure 1:**
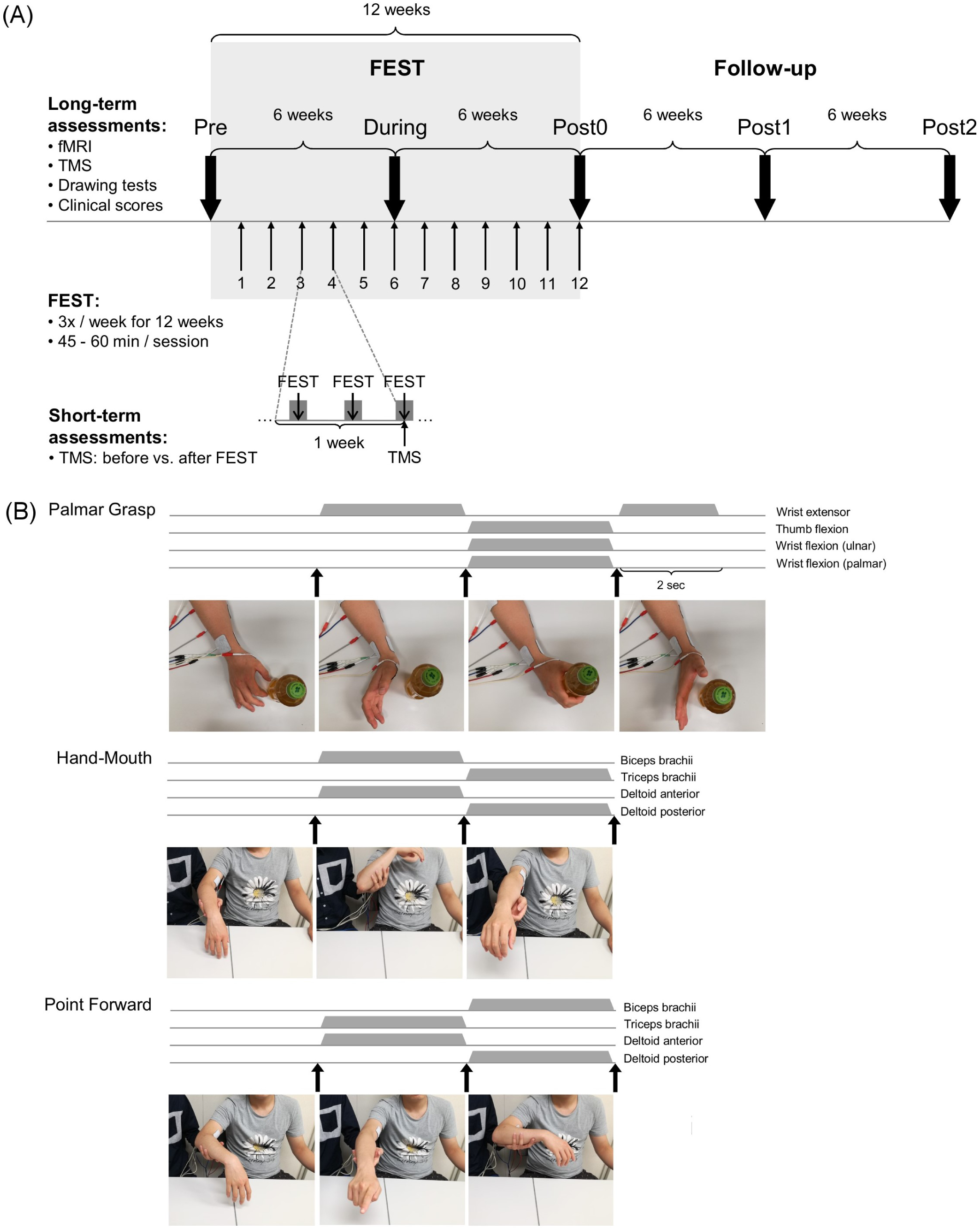
Experimental setup. **(A)** Experimental protocol - Functional electrical stimulation therapy (FEST) was delivered over the course of 12-weeks with three sessions per week and each session lasting 45-60 min. Long-term assessments were carried out at baseline (Pre), after 6-weeks and 12-weeks of FEST (During and Post0), as well as during follow-up 6-weeks and 12-weeks after FEST (Post1 and Post2) and they included: functional magnetic resonance imaging (fMRI), transcranial magnetic stimulation (TMS), drawing tests, and clinical test evaluations. Short-term assessments were carried out once per week over the course of 12-weeks to compare before and after each FEST session using TMS assessments; **(B)** Each FEST training session consisted of three functional training protocols including the palmar grasp - to generate hand opening, hand-mouth - to generate elbow and shoulder flexion, and point forward - to generate hand pointing forward, by activating a sequence of muscles activations.

### 2.3. Assessment protocols

Timeline of assessments is summarized in Figure 1A. Assessments were carried out to evaluate cortical and corticospinal circuits associated with upper-limbs as well as upper-limb functional performance and clinical scores. Short-term cortical changes were assessed once per week over the course of 12-weeks of training immediately before and after each FEST session using transcranial magnetic stimulation (TMS). Long-term assessments were carried out every 6-weeks over the course of the 12-weeks of FEST and during the 12-weeks follow-up after the intervention was complete. Specifically, long-term changes were assessed before the training at baseline (Pre), after 6-weeks of the training (During), and immediately after 12-weeks of FEST (Post0), as well as 6-weeks after FEST was completed (Post1) and 12-weeks after FEST was completed (Post2). Long-term cortical changes and corticospinal excitability were evaluated using TMS and fMRI, while functional performance was assessed using an instrumented drawing test and clinical scores.

#### 2.3.1. Transcranial magnetic stimulation (TMS)

TMS sessions were carried out during both short-term and long-term assessments. During the assessments, participant remained seated comfortably on the chair with the right hand supported on the table. Electromyographic (EMG) activities were recorded using bipolar Ag/AgCl surface electrodes (Vitrode F-150S, Nihon Koden, Tokyo, Japan) from the right (intervention) hand: (i) first dorsal interosseous (FDI) and (ii) abductor pollicis brevis (APB) muscles. A ground electrode was placed on the elbow of the right arm. It was ensured that the EMG electrodes were placed roughly on the same locations of the muscle between assessment days. EMG signals were band-pass filtered (15-1,000 Hz), amplified (1,000×; MEG-6108, Nihon Koden, Tokyo, Japan) and sampled at 4,000 Hz using an analog-to-digital converter (Powerlab/16SP, AD Instruments, Castle Hill, Australia).

Using a mono-phasic magnetic stimulator (Magstim 200, Magstim Co., Whitland, UK) through a figure-of-eight coil, single-pulse TMS was delivered over the left primary motor cortex (M1) area that was optimal for inducing MEPs in the right FDI. The “hot spot” location was determined and defined with respect to cranial landmark as references during the baseline assessment (Pre). The same “hot spot” location was used as a for all subsequent assessments (During, Post0, Post1, and Post2). The MEPs were always evoked with the participant keeping voluntary contraction at 10% maximal voluntary contraction (MVC) effort of the FDI muscle during the finger pinch task since there were no visible MEP responses at rest during baseline assessments (Pre). Contractions were maintained by holding a force sensor (OKLU-100K-S1-H18, Frontier Medic, Hokkaido, Japan) with his right thumb and index fingers, while the force level was shown on a visual display. The motor threshold (MT) for evoking MEPs was the minimum TMS intensity to elicit peak-to-peak amplitudes of at least 50 μV from the FDI muscle in five of ten consecutive trials (Groppa et al. 2012). It was ensured that the MEPs of the APB muscle could also be evoked and recorded simultaneously.

During short-term and long-term assessments, the input-output relationship between TMS stimulation intensity and MEP responses amplitude was obtained by applying TMS at 60, 70, 80, 90 and 100% of the TMS stimulator intensity. Three trials were performed at each TMS intensity and the responses obtained for each muscle (FDI and APB) at each intensity (Ridding et al. 2001). Since MEPs were recorded during active contractions at 10% MVC, it was also possible to record the CSP of the MEPs from the same trials. Three CSP trials were also calculated from the responses evoked at 70% of the stimulator output (Farzan 2014).

During long-term assessments, MEP maps of corticospinal responses of each muscle were recorded by applying TMS at 70% of the stimulation output, which was determined to be the 120% MT stimulation intensity during the baseline (Pre) assessment and remained unchanged. During each assessment, the participant was asked to keep voluntary contractions at 10% of MVC of the FDI muscle. The MEP map was centered at the FDI “hot spot” location, which was defined with respect to cranial landmark during the baseline (Pre) assessment and remained unchanged. The MEP map was then expanded to the surrounding points on the 10×10 cm grid with a 1 cm resolution (100 cm^2^ area) around the “hot spot” location using pre-determined markings on a tight-fitting cap. Three stimuli were delivered at each location in a semi-randomized order at a rate of approximately every 6 sec and averaged to obtain a peak-to-peak amplitude response for each location (Mortifee et al. 1994; Ridding et al. 2001).

#### 2.3.2. Functional magnetic resonance imaging (fMRI)

During fMRI sessions, which were carried out during long-term assessments, the participant remained in the supine position in an MRI scanner (MAGNETOM Plisma, Siemens, Germany) and was asked to perform: (i) hand grip; and (ii) finger pinch force matching tasks with the right (intervention) hand, while holding a force sensor (OKLU-100K-S1-H18, Frontier Medic, Hokkaido, Japan). The force matching tasks was a trapezoidal pursuit consisting of four phases: rest, ascending, keep, and descending, each lasting 10 s. The target force level (keep phase) was set to 20% of the MVC effort (Ward et al. 2003), while the ascending and descending phase linearly increased and decreased to the target force over the course of 10 s. The participant could see the target force on the visual display, which they attempted to match during the experimental trials. The task was repeated a total of four times during each assessment. The MVC levels were determined prior to the experiment for the hand grip and finger pinch tasks. During fMRI assessments, the participant was asked to follow the target force trajectories as precisely as possible. All MRI images were acquired using a 3T MRI scanner (MAGNETOM Plisma, Siemens, Germany). Functional T2*-weighted echo-planar images that reflect blood oxygenation level-dependent (BOLD) responses (Ogawa et al. 1990) as well as high-resolution T1-weighted structural images were collected (see Supplementary information: fMRI data acquisition).

#### 2.3.3. Drawing tests

To evaluate upper-limb fine motor function, which was carried out during long-term assessments, the participant was asked to perform: (i) tracing and (ii) target tracking tasks of a sine wave (wavelength: 50 mm, amplitude: 25 mm, distance: 150 mm) using an instrumented tablet system (TraceCoder® Version 1.0.8, Surface Pro4, SystemNetwork, Osaka, Japan) (Itotani et al. 2016). During the assessments, the participant was comfortably seated in a chair with his elbow on the table and flexed at 90°. During the tracing task, the participant was instructed to follow the outline of a sine wave at his preferred speed without a moving target, while during the target tracking task, the participant was instructed to follow the moving target on the tablet screen which moved on a sine wave at 12 mm/sec. For both tasks, the participant was asked to draw as precisely as possible. Two trials, each consisting of three sine waves, were recorded for each of the tracing and tacking tasks. Before each assessment, a practice period was allowed.

#### 2.3.4. Clinical assessments

Clinical scores, which were evaluated during long-term assessments, included functional independence measure (FIM) (Granger and Hamilton 1992), Fugl-Meyer assessment (FMA) (Fugl-Meyer 1980), and Motor Activity Log (MAL) (van der Lee et al. 2004). All tests were performed by the same trained therapist.

### 2.4. Data analysis

#### 2.4.1. MEPs

All MEP analysis was performed using a custom program written in Matlab (The MathWorks Inc., USA). To evaluate the input-output curve relationship between the TMS stimulation intensity and the MEP responses for the FDI and APB muscles, MEP peak-to-peak amplitudes of each muscle for each of the three repeated trials at each stimulation intensity (60, 70, 80, 90, and 100% of the TMS stimulator output) were first calculated. The MEP amplitudes were plotted relative to the TMS stimulation intensity and a linear fit line was obtained using simple linear regression. The slope of the linear regression line was used to define the three repeated trials gain parameters of the input-output relationship curve (Farzan 2014).

The CSP duration was defined for each muscle for three repeated trials as the time between the end of the MEP (i.e., where EMG activity was below 3SD of mean pre-stimulus activity) and the time at which the post-stimulus EMG returned to the pre-stimulus EMG activity (i.e., where EMG activity exceeded 3SD of the mean pre-stimulus activity) (Farzan 2014).

Corticospinal representation MEP maps were calculated from the MEP peak-to-peak amplitudes of each point on the 100 cm^2^ area (10×10 cm map with 1 cm resolution). The three repeated trials for each point were first averaged and normalized with the peak MEP amplitude on the map for each assessment day. The MEP map was then constructed from the average MEP amplitudes from each point on 10×10 cm grid using Matlab’s ‘gridfit’ function to define 2,500 partitions within 100 cm^2^ area (D’Errico 2005). Finally, activated area on the 100 cm^2^ map was calculated by taking the ratio of the number of partitions where the approximated MEP exceeded 10% of maximum MEP (aMEP_10%_) relative to all partitions (N_total_ = 2,500): 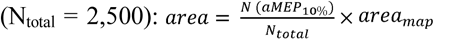, where area_map_ is 100 cm^2^ (van de Ruit et al. 2015).

#### 2.4.2. fMRI

All fMRI data analysis was performed using Statistical Parametric Mapping (SPM12, Wellcome Trust Center for Neuroimaging, London, UK) software implemented in Matlab (The MathWorks Inc., USA). First, data preprocessing procedures were applied (see Supplementary information: fMRI data processing). After the preprocessing, the general linear model regression to the time course data was obtained to estimate the amount of neural activation (Friston et al. 1994; Friston et al. 1995). Whole brain analysis was performed to depict the general features of brain activations during the hand grip and finger pinch tasks. First, the brain regions where the BOLD signals increased during the hand grip and finger pinch were depicted by evaluating the T-values obtained from each session to contrast a task specific voxel by voxel activation map. The threshold was set at voxel level p<.001 (uncorrected) and cluster level p<.050 family-wise error correction (FWE) (Woo et al. 2014).

Next, the region of interest (ROI) was set in six anatomical hand areas defined bilaterally: primary motor cortex (M1: *x*=±37, *y*=−21, *z*=58) (Mayka et al. 2006), sensory cortex (S1: *x*=± 40, *y*=−24, *z*=50) (Mayka et al. 2006), secondary somatosensory cortex (S2: *x*=±58, *y*=−27, *z*=30) (Iftime-Nielsen et al. 2012), parietal rostroventral area (PR: *x*=±54, *y*=−13, *z*=19) (Hinkley et al. 2007), supplementary motor area (SMA: *x*=±20, *y*=−8, *z*=64) (Ciccarelli et al. 2006), premotor cortex (PM: *x*=±8, *y*=−6, *z*=64) (Ciccarelli et al. 2006). These ROI regions were chosen based on the previous studies that investigated cortical effects of FES (Blickenstorfer et al. 2009; Gandolla et al. 2016). In addition, the most activated voxel (peak voxel) in the contralateral M1 region was calculated to define the most active ROI location (Verstynen et al. 2005). A control region was defined as the hippocampus gyrus (HC: *x*=−22, *y*=−34, *z*=−8 for contralateral and *x*=32, *y*=−30, *z*=−8 for ipsilateral) (Hayes et al. 2011), which was not associated with hand movements. The BOLD signal time-series data from all ROIs was extracted and calculated as the percent signal change for each force matching phase volume (ascending, keep, and descending) relative to the mean BOLD signal in the rest phase volume (Uehara et al. 2019). The task was repeated four times, resulting in twelve measurements for each assessment point.

#### 2.4.3. Drawing tests

Tracing and target tracking tasks were evaluated using the following parameters to assess performance: (i) sum of error - difference between the target coordinates of the sine wave and participant’s pen in in the x direction (medio-lateral), y direction (antero-posterior), and xy direction (sum of squared error); (ii) velocity - mean velocity during the tasks; (iv) acceleration - mean acceleration during the tasks; and (iv) pressure - mean pressure exerted during the tasks. The parameters were calculated for each full sine wave and the task was repeated two times, resulting in six measurements for the tracing and sine wave tracking tasks for each assessment point. All parameters were calculated using a custom program written in Matlab (The MathWorks Inc., USA).

#### 2.4.4. Clinical assessments

Clinical scores for the FIM, FMA, and MAL tests were tabulated and evaluated by a trained occupational therapist and compared between different assessment points.

### 2.5. Statistics

Short-term TMS assessments were analyzed using paired samples t-test to compare the input-output curve slope and CSP between assessment points (before and after). Long-term TMS assessments were analyzed using the one-way repeated measures analysis of variance (ANOVA) to compare the input-output curve slope and CSP between assessment points (Pre, During, Post0, Post1, and Post2). Same statistical procedures were applied to compare long-term fMRI cortical activations during hand grip and finger pinch tasks in the peak activated voxel in M1 as well as in the contralateral and ipsilateral hemisphere in each ROI (M1, S1, S2, PR, SMA, PM, and HC), as well as drawing task error (x, y, and xy directions), velocity, acceleration, and pressure between assessment points. When significant results were found on the ANOVA, post-hoc multiple comparisons with Holm adjustment were conducted to compare Pre to other assessment points. Parametric tests were chosen since the Shapiro-Wilk test was used to confirm that most data were normally distributed. Short-term assessments were performed before and after each FEST session over the 12-weeks, while long-term assessments were performed on repeated trials on each assessment point. Statistical comparisons were performed using SPSS Statistics (IBM Corp., Armonk, NY, USA). Significance level for all tests was set to p<.050.

## 3. Results

### 3.1. Short-term effects

Short-term TMS assessment comparisons are summarized in Figures 2A and 2B. Input-output curve showed no statistically significant differences between slopes of FDI (t_(11)_=-2.137, p=.056) and APB (t_(10)_=0.226, p=.830) muscles after each FEST session, compared to before the session (Figure 2A). However, CSP showed statistically significant decrease in the silent period in both FDI (t_(11)_=2.503, p=.002) and APB (t_(10)_=4.000, p=.029) muscles after each FEST session, compared to before the session (Figure 2B).

**Figure 2:**
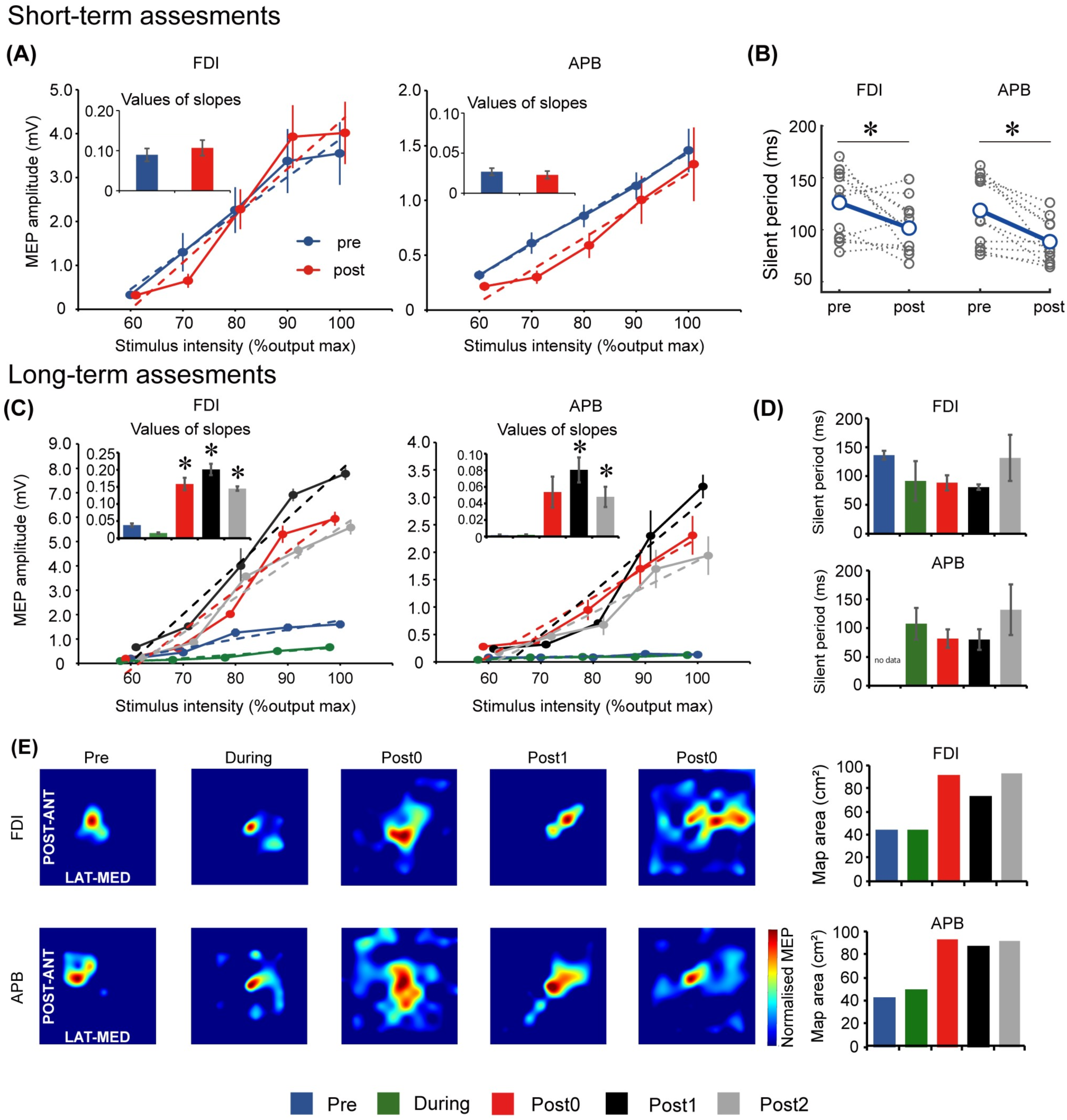
Motor evoked potential (MEP) results for the short-term assessments. **(A)** Input-output relationship curve for the first dorsal interosseous (FDI) and abductor pollicis brevis (APB) muscles. Dotted lines indicate simple linear regression lines of the curves before and after one functional electrical stimulation therapy (FEST) session. Bar graphs indicate values of regression line slope and standard error; **(B)** Cortical silent period (CSP) for the FDI and APB muscles before and after one FEST session. Gray dotted lines indicate data of each day. ***MEP results for the long-term assessments*** *-* **(C)** Input-output relationship curve for the FDI and APB muscles. Dotted lines indicate simple linear regression lines of the curves at baseline (Pre), after 6-weeks and 12-weeks of FEST (During and Post0) as we as during follow-up assessments 6-weeks and 12-weeks after FEST (Post1 and Post2). Bar graphs indicate values of regression line slope and standard error; **(D)** CSP for the FDI and APB muscles during Pre, During, Post0, Post1 and Post2 assessments. Bar graphs indicate values of regression line slope and standard error; **(E)** MEP maps before and after FEST for the FDI and APB muscles. The size of the MEP activated is approximated by the heatmap color scale, which denotes amplitudes normalized to the maximum value in assessment. Bar graphs indicate the calculated area of the MEP map. Legend: * p<.050.

### 3.2. Long-term effects

#### 3.2.1. TMS

Long-term TMS assessment comparisons are summarized in Figures 2C, 2D, and 2E. Input-output curve showed statistically significant differences between assessment points in both FDI (F_(4,8)_=147.678, p<.001) and APB (F_(4,8)_=31.790, p<.001) muscles. Post hoc comparisons (Figure 2C) showed that the slope increased significantly after 12-weeks of FEST (Post0) in the APB muscle and that it remained for at least another 12-weeks after the FEST intervention was completed (Post1 and Post2) in both FDI and APB muscles. CSP showed that there were no statistically significant differences between assessment points in both FDI (F_(4,8)_=3.001, p=.086) and APB (F_(3,6)_=2.261, p=.182) muscles (Figure 2D). Finally, descriptive comparisons of MEP maps suggest that the area in the motor cortex in both FDI and APB muscles increased after 12-weeks of FEST (Post0) and that it remained for at least another 12-weeks after the FEST intervention was completed (Post1 and Post2) in both FDI and APB muscles (Figure 2E).

#### 3.2.2. fMRI

Long-term assessment fMRI activations of the whole brain during the hand grip task are summarized in Figure 3A. Peak activated voxel in M1 showed statistically significant differences between assessment points for the hand grip task (F_(4,44)_=5.814, p=.001). Post hoc comparisons (Figure 3A) showed that activation significantly increased after 12-weeks of FEST (Post0) but returned to baseline after the FEST intervention was completed (Post1 and Post2). ROI analysis for the hand grip task is summarized in Figure 3B. Contralateral hemisphere comparisons showed that activations in M1 (F_(4,44)_=6.070, p=.001), PR (F_(4,44)_=7.113, p<.001), SMA (F_(4,44)_=7.064, p<.001), and PM (F_(4,44)_=144.163, p<.001) had statistically significant differences, while S1 (F_(4,44)_=3.781, p=.010; note: no statistically significant post hoc comparisons were shown), S2 (F_(4,44)_=2.485, p=.057), and HC (F_(4,44)_=0.256, p=.905) had no significant differences between assessment points. Post hoc comparisons (Figure 3B top) indicate that contralateral motor related areas (M1, PR, SMA and PM) primarily increased activations after 12-weeks of FEST (Post0) during the hand grip task. Ipsilateral hemisphere comparisons showed that activations in M1 (F_(4,44)_=6.538, p=.001) and S1 (F_(4,44)_=3.925, p=.008) had small statistically significant differences, while S2 (F_(4,44)_=0.835, p=.510), PR (F_(4,44)_=0.224, p=.925), SMA (F_(4,44)_=1.275, p=.294), PM (F_(4,44)_=1.029, p=.403), and HC (F_(4,44)_=0.545, p=.704) had no significant differences between assessment points. Post hoc comparisons (Figure 3B bottom) indicate little or not ipsilateral activations during the hand grip task.

**Figure 3:**
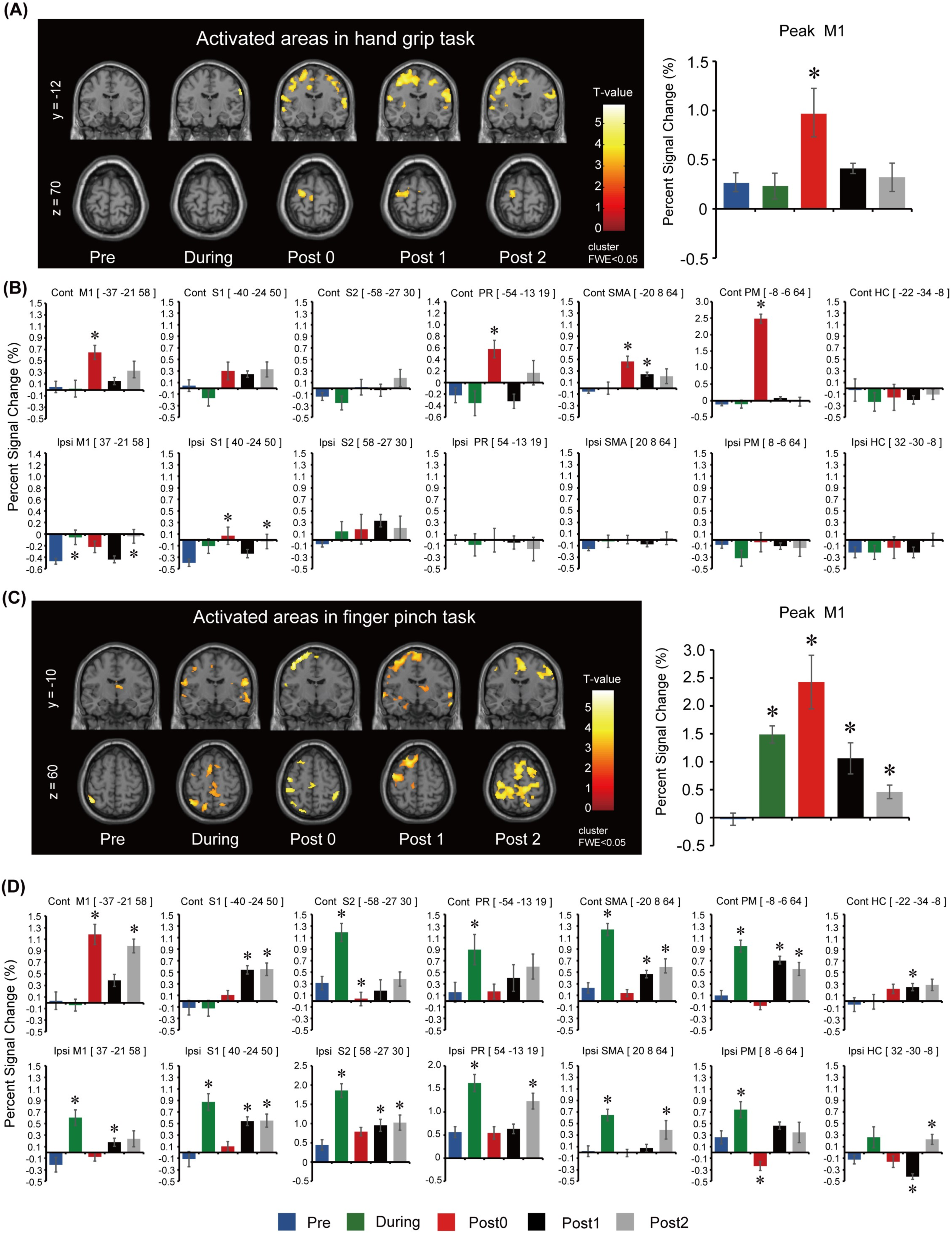
Functional magnetic resonance imaging (fMRI) results for the long-term assessments during the hand grip task. **(A)** Activated regions during right (intervention) hand grip task. To observe the whole brain activity, the coordinates of y=−12 and z=70 planes were used. T-values are plotted, and the threshold was set at voxel level p<.001 (uncorrected) and cluster level p<.050 (FWE). Assessments were carried out at baseline (Pre), after 6-weeks and 12-weeks of FEST (During and Post0), as well as during follow-up assessments 6-weeks and 12-weeks after FEST (Post1 and Post2). Region of interest (ROI) results of the most activated voxel in the primary motor cortex (M1) for each assessment are shown next to the activated regions; **(B)** ROI results based on anatomical regions in the M1 as well as the sensory cortex (S1), secondary somatosensory cortex (S2), parietal rostroventral area (PR), supplementary motor area (SMA), premotor cortex (PM), and the hippocampus gyrus (HC). The upper bar graphs show activity of the contralateral hemisphere (Contra) and the lower bar graphs shows activity of the ipsilateral hemisphere (Ipsi). ***fMRI during the finger pinch task*** *-* **(C)** Activated regions during right (intervention) finger pinch task. To observe the whole brain activity, the coordinates of y=−10 and z=60 planes were used. T-values are plotted and the threshold was set at voxel level p<.001 (uncorrected) and cluster level p<.05 (FWE). Assessments were carried out at Pre, During, Post0, as we as Post1 and Post2. ROI results of the most activated voxel in the primary motor cortex (M1) for each assessment were shown next to the activated regions; **(D)** ROI results based on anatomical regions in the M1 as well as S1, S2, PR, SMA, PM, and HC. The upper bar graphs show activity of the contralateral hemisphere (Contra) and the lower bar graphs shows activity of the ipsilateral hemisphere (Ipsi).

Long-term assessment fMRI activations of the whole brain during the finger pinch task are summarized in Figure 3C. Peak activated voxel in M1 showed statistically significant differences between assessment points for the finger pinch task (F_(4,44)_=13.319, p<.001). Post hoc comparisons (Figure 3C) showed that activation significantly increased after 6-weeks and 12-weeks of FEST (During and Post0) as well as in the 6-week and 12-week follow-up period (Post 1 and Post2). ROI analysis for the finger pinch task is summarized in Figure 3D. Contralateral hemisphere comparisons showed that activations in M1 (F_(4,44)_=21.505, p<.001), S1 (F_(4,44)_=10.306, p<.001), S2 (F_(4,44)_=19.246, p<.001), PR (F_(4,44)_=4.471, p=.004), SMA (F_(4,44)_=29.309, p<.001), PM (F_(4,44)_=24.644, p<.001), as well as HC (F_(4,44)_=3.308, p=.019) all had statistically significant differences between assessment points. Post hoc comparisons (Figure 3D top) indicate contralateral motor cortex activations (M1) increased after 12-weeks of FEST (Post0) as well as widespread changes in all other areas after 6-weeks of FEST (During) which persisted in follow-up (Post1 and Post2) during the finger pinch task. Ipsilateral hemisphere comparisons showed that activations in M1 (F_(4,44)_=9.227, p<.001), S1 (F_(4,44)_=3.925, p=.008), S2 (F_(4,44)_=17.585, p<.001), PR (F_(4,44)_=11.634, p<.001), SMA (F_(4,44)_=11.516, p<.001), PM (F_(4,44)_=11.587, p<.001), as well as HC (F_(4,44)_=9.004, p<.001) all had statistically significant differences between assessment points. Post hoc comparisons (Figure 3D bottom) indicate widespread changes in all areas after 6-weeks of FEST (During) which persisted in follow-up (Post1 and Post2) during the finer pinch task.

#### 3.2.3. Drawing tests

Long-term assessment drawing test comparisons are summarized in Figure 4. Tracing task comparisons showed that velocity (F_(4,20)_=5.219, p=.005), acceleration (F_(4,20)_= 4.333, p=.011), and pressure (F_(4,20)_=10.361, p<.001) had statistically significant differences, while sum of x errors (F_(4,20)_=1.710, p=.187), sum of y errors (F_(4,20)_=2.432, p=.081), and sum of xy errors (F_(4,20)_=1.885, p=.152) had no significant differences between assessment points. Post hoc comparisons (Figure 4C top) indicate decreased velocity and acceleration after 12-weeks of FEST (Post0) which persisted in follow-up (Post1 and Post2) during the tracing task (note: pressure also seemed to decrease in all time points except Post2), as well as a similar trend in error reduction, although not statistically significant.

**Figure 4:**
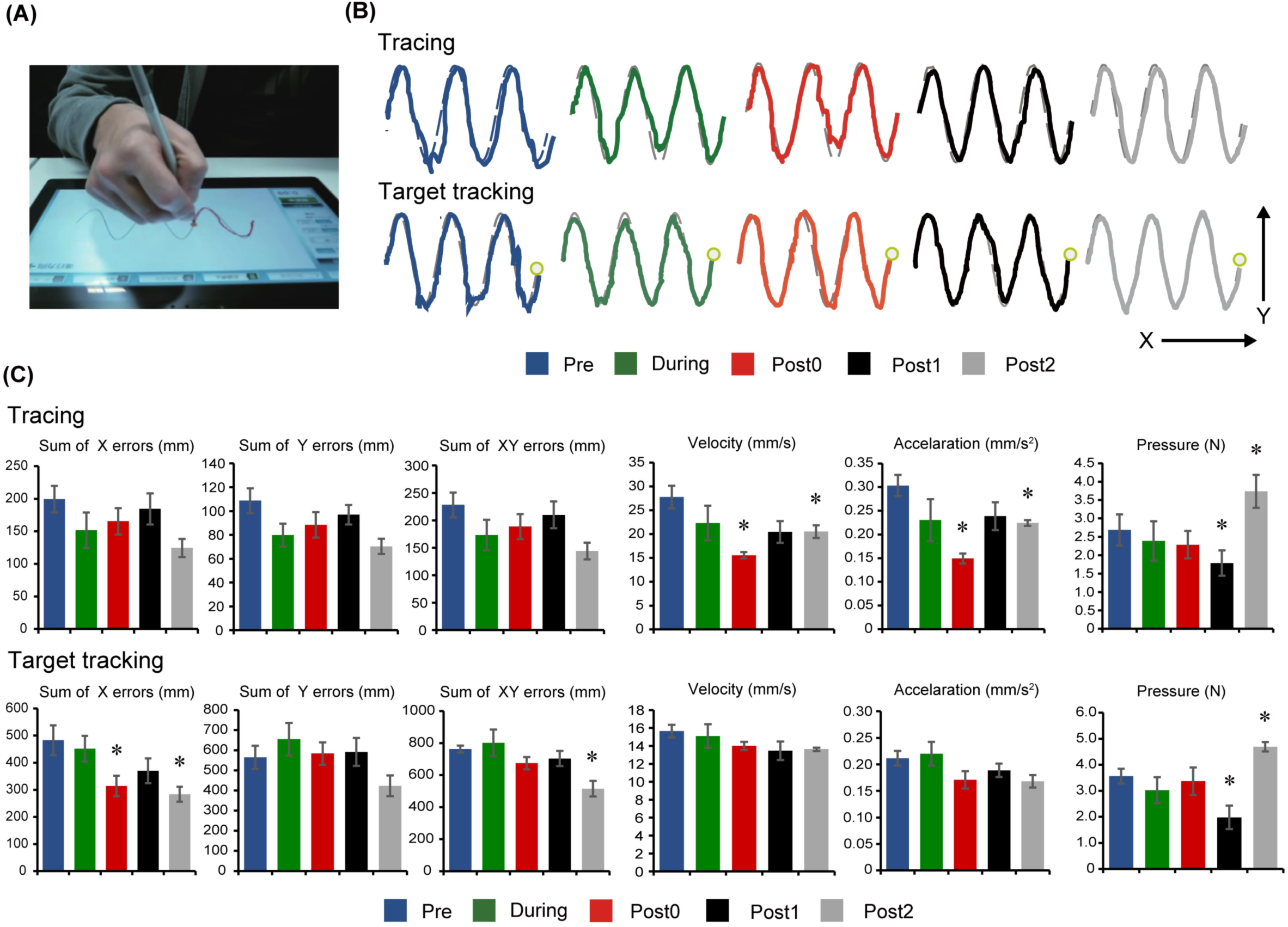
Drawing test results. **(A)** Experimental setup showing the instrumented tablet with the participant; **(B)** Representations of the participant’s performances on the drawing tests at baseline (Pre), after 6-weeks and 12-weeks of FEST (During and Post0), as we as during follow-up assessments 6-weeks and 12-weeks after FEST (Post1 and Post2) are shown. Tracing performance is shown in the upper graphs, when the participant was required to follow the outline of a sine wave at a self-selected speed. Target tracking performance is shown in the lower traces, when the participant was required to follow a moving target on the screen; **(C)** The sum of error (x, y, and xy directions), velocity, acceleration, and pressure performance during the tracing task are shown in the upper graphs and the target tracking task in the lower graphs.

Target tracking task comparisons showed that sum of x errors (F_(4,20)_=3.887, p=.017), sum of xy errors (F_(4,20)_=4.570, p=.009), and pressure (F_(4,20)_=5.727, p<.001) had statistically significant differences, while sum of y errors (F_(4,20)_=2.290, p=.095), velocity (F_(4,20)_=1.232, p=.329), and acceleration (F_(4,20)_=2.106, p=.118) had no significant differences between assessment points. Post hoc comparisons (Figure 4C bottom) indicate decreased error predominantly in the medio-lateral x-direction (note: pressure also seemed to decrease in all time points except Post2).

#### 3.2.4. Clinical assessments

Long-term clinical score results are summarized in Table 1. The FIM and FMA scores were not different after 6-weeks (During) and 12-weeks (Post0) of FEST, as well as during the follow-up assessments at 6-weeks (Post1) and 12-weeks (Post2) after the FEST intervention was completed, compared to baseline (Pre). However, the MAL score increased by 1 point after 6-weeks of FEST (During) and remained after 12-weeks of FEST (Post0) and for at least another 12-weeks after the FEST intervention was completed (Post 1 and Post 2) (Table 1).

**Table 1:** Clinical measurements scores, including the functional independence measure (FIM) self-care, Fugl-Meyer assessment (FMA) of the upper-limb (U/L) function and Motor Activity Log (MAL) amount of use score (AS) and how well score (HW).

|  | Pre | During | Post0 | Post1 | Post2 |
| --- | --- | --- | --- | --- | --- |
| **FIM self-care** (max score: 42) | 42 | 42 | 42 | 42 | 42 |
| **FMA U/L** (max score: 66) | 63 | 63 | 63 | 63 | 63 |
| **MAL AS and HW** (max score: 150/150) | 78/92 | 79/92 | 79/92 | 79/92 | 79/92 |

## 4. Discussion

### 4.1. Evidence of cortical re-organization after FEST

Our results showed the time course of cortical re-organization elicited by a FEST intervention in an individual with chronic TBI. Specifically, short-term assessment results showed reduced cortical silent period (Figure 2B). Cortical silent period refers to an interruption of voluntary muscle activity by TMS applied over the contralateral motor cortex (Wolters et al. 2008; Farzan 2014). It is generally agreed that spinal inhibitory mechanisms contribute to the silent period up to its first 50 ms, while the later part is generated exclusively by inhibition within the motor cortex (Wolters et al. 2008). It has previously been shown that FES can inhibit spinal reflex excitability (Kawashima et al. 2013). Moreover, consistent to our results, electrical stimulation of cutaneous nerves in the upper-limbs was also shown to shorten the cortical silent period (Hess et al. 1999; Classen et al. 2000), which suggests involvement of cortical-level sensorimotor integration (Wolters et al. 2008). Cutaneous and afferent feedback from FEST may activate the somatosensory cortex, which may affect cortico-cortical connections (Carson and Buick 2019). It has previously been demonstrated that somatosensory cortices are activated during electrical stimulation of muscles (Korvenoja et al. 1999; Boakye et al. 2000; Nihashi et al. 2005). In fact, our fMRI results also showed an increase in signal intensity not only in M1 but also in S1 and S2 during long-term assessments after FEST, which supports these considerations (Figures 3B and 3D). Therefore, short-term effects of FEST are likely related to sensorimotor integration through intracortical inhibition or possibly spinal reflex inhibition after each FEST session.

Our long-term assessment results indicate that the slope of MEP input-output curve was not facilitated after 6-weeks of FEST, while there was significant facilitation after 12-weeks, which remained even during follow-up (Figure 2C). The slope of the MEP input-output curve reflects the strength of corticospinal projections to the target muscles (Farzan 2014) and can become less steep with GABA_A_ (inhibitory) receptor agonist (lorazepam), while administration of an indirect dopaminergic-adrenergic (excitatory) agonist (D-amphetamine) increased the slope (Boroojerdi et al. 2001). Taken together, our results indicate considerable long-term facilitation of corticospinal excitability after 12-weeks of FEST which may persist for another 12-weeks even in the absence of any intervention in an individual with TBI, possibly via upregulation of dopaminergic excitatory receptors and/or downregulation of GABAergic inhibitory receptors.

Increased corticospinal excitability can likely be explained by larger area over which MEPs can be obtained in the hand muscles, which were shown in our study. Specifically, MEP map results indicate enlarged hand muscle representations within the M1 after 12-weeks of FEST and during follow-up (Figure 2E). Motor maps obtained using TMS-evoked MEPs are reliable for extracting useful somatotopic information from the primary motor cortex (Wilson et al. 1993; Wassermann et al. 1992). It was previously shown that 2-hours of electrical nerve stimulation can produce larger areas over which MEPs can be evoked (Ridding et al. 2001). We confirmed considerable expansion of the motor areas which are consistent with the time-course of changes of MEP facilitation evoked over a single “hot spot” location during long-term follow-ups. While motor evoked responses could reflect cortical and/or spinal excitability, increased motor map area and subsequent MEP amplitude facilitation (Ridding and Rothwell 1997) confirm cortical re-organization after FEST in an individual with chronic TBI in our study.

Cortical re-organization was further corroborated by our fMRI data, which showed larger BOLD responses after 12-weeks of FEST compared to baseline assessments during both hand grip and finger pinch tasks (Figures 3A and 3C). Peak signal intensity within the M1 during the hand grip task was significantly increased after 12-weeks of FEST, while it returned to baseline during follow-up (Figure 3A). On the other hand, during the finger pinch task, the peak M1 signal was significantly increased after 6-weeks and 12-weeks of FEST as well as during follow-up assessments, while a gradual reduction of signal compared to after the intervention was observed when FEST was completed (Figure 1C). The time course of cortical changes obtained using fMRI in the contralateral M1 ROI (Figures 1B and 1D) is consistent to the MEP results obtained using TMS. Analysis of other ROI voxels indicates widespread changes not only in the M1, but also in the parietal rostroventral area (PR), supplementary motor area (SMA), and premotor cortex (PM) area during both hand grip and finger pinch tasks. During the finger pinch task, which is a fine motor skill that was most notably impaired in our participant, the primary (S1) and secondary somatosensory cortex (S2) changes were also shown, as well as overall earlier activations (i.e., 6-weeks after FEST) and more widespread changes in both contralateral and ipsilateral hemispheres which included the control region (HC) that was not expected to change. Evidence from various neuroimaging studies has previously shown that somatosensory cortices, including both S1 and S2 areas, are activated during electrical stimulation of muscles (Korvenoja et al. 1999; Boakye et al. 2000; Nihashi et al. 2005). When FES is applied at motor threshold intensity to generate flexion and extension wrist movements, cortical activations in the contralateral M1, S1 and PM areas, as well as bilateral S2 and SMA activation were shown to be activated (Blickenstorfer et al. 2009). During FEST, the participant was asked to attempt each movement before the therapist applied FES to activate the appropriate muscles. Long-term repeated sensorimotor integration facilitated by FES during task-specific upper-limb training that includes voluntary engagement may therefore elicit cortical re-organization. Specifically, integration of motor commands during voluntary movement attempt and sensorimotor network activation through FES are the candidate mechanisms of long-term cortical changes after FEST. Intact motor areas topologically adjacent to the damaged site within the M1 and areas outside of motor cortex may assume control over the affected muscles via intracortical connectivity networks after task-specific repetitive training by Hebbian synaptic strengthening (Weiller et al. 1992; Seitz et al. 2005; Nudo et al. 2013). Our findings therefore indicate that widespread cortical re-organization caused by FEST can elicit neuroplasticity after chronic TBI in cortical areas related to fine motor function.

### 4.2. Carry-over effects after FEST

Consistent to our results that demonstrated carry-over effects during follow-up assessments (Figures 2 and 3), other evidence also points that sustained cortical changes can outlast the intervention period. Therapeutic applications of FES delivered over longer periods indicated long-term cortical re-organization after the intervention (Shin et al. 2008; Sasaki et al. 2012). Specifically, 30 min of FES-assisted finger flexion and extension applied once per day for a total of 12-weeks was shown to elicit cortical changes in the somatosensory cortex after the intervention, which were correlated to the improvements in the motor function in chronic hemiplegia patients (Sasaki et al. 2012). Similarly, 60 min of FES wrist extension applied 5 days per week for a total of 10-weeks resulted in shifting of the somatosensory area activations from ipsilateral to the contralateral hemisphere after the intervention, which was related to significant improvements in the motor function in chronic stroke patients (Shin et al. 2008). Taken together, our results suggest that approximately 40-hours of task-specific and repetitive FEST are required to induce cortical re-organization associated with the upper-limb control (Shin et al. 2008; Sasaki et al. 2012), while only some changes were observed with less training after 6-weeks of FEST (Figures 3C and 3D). Importantly, our current study also demonstrated that long-term cortical re-organization could persist for several months (i.e., for as long as 12-weeks) after FEST, which is consistent with clinical recovery profiles shown by our group (Kapadia et al. 2011). Considering that the individual in our current study was in the chronic stage (>7 years) after the injury, spontaneous recovery can be ruled out. Evidence therefore suggests that cortical re-organization after TBI can be elicited using FEST and that carry-over effects may outlast the intervention period.

### 4.3. Fine motor function improvements after FEST

Clinical scores suggest that the individual in our study had a relatively high level of motor function at the onset of the FEST intervention. Specifically, our participant had a FIM score of 42/42 (Table 1), which indicates complete independence in activities of daily living, including motor scores, communication, and social cognition (Granger and Hamilton 1992). Similarly, the upper-limb portion of the Fugl-Meyer assessment was 63/66 (Table 1), indicating high level of upper-limb function. As expected, neither the FIM nor the Fugl-Meyer assessment scores were changes after FEST. While the MAL score increase from 78 to 79/92 after 6-weeks of FEST (Table 1) may indicate minimal clinically important improvements (Simpson and Eng 2013), no major changes in gross motor function were shown due to ceiling effects.

However, drawing test results showed fine motor function improvements after FEST (Figure 4C). Specifically, the tracing task, which required following the outline of a sine wave at a self-selected speed, showed significantly decreased mean velocity and acceleration after 12-weeks of FEST and during follow-up, which may suggest less abrupt and smoother movements during the target tracing task (Figure 4C top). While the error seemed to decrease during both tasks, significant reduction during the target tracking task, which required following a moving target on the screen, was shown after 12-weeks of FEST and during follow-up, indicating improved fine motor function performance (Figure 4C bottom). It has been suggested that cortical changes resulting from FES interventions or other rehabilitation programs are not always correlated to improvements in motors function (Quandt and Hummel 2014), or that motor function can event initially deteriorate (Murata et al. 2008). Nonetheless, our results showed changes on the drawing tests after FEST which are indicative of improved performance. Similarly, improved tracing task performance was shown after 4-weeks of upper-limb FEST in a clinical randomized trial in individuals with hemiplegia (Popovic et al. 2003). More intense FEST protocols also improved drawing performance and were associated with reduced spasticity after stroke (Kawashima et al. 2013). Similarly, improvements in drawing accuracy were also reported in individuals with chronic stroke after 10-weeks of FES upper-limb therapy, consistent to increased cortical activations, while the control group which did not display altered cortical activations also did not improve on the drawing test (Shin et al. 2008). Electrical stimulation may therefore elicit cortical re-organization, which can ultimately serve as a basis for improved functional capacity (Traversa et al. 1997; Fraser et al. 2002; Carson and Buick 2019). Our current study utilized the FEST protocols developed by our group, which were shown in randomized clinical trials to improve motor function after neurological injuries (Thrasher et al. 2008; Kapadia et al. 2011). Using these protocols, we demonstrated considerable cortical re-organization, which was accompanied by improved fine motor function after FEST in an individual with chronic TBI.

### 4.4. Limitations

A limitation of this study is the small sample size and lack of a control group to examine benefits of equivalent conventional therapy. Our team has previously demonstrated in randomized controlled clinical trials that upper-limb FEST intervention is superior for improving hand motor function compared to conventional therapy after stroke and incomplete SCI (Thrasher et al. 2008; Kapadia et al. 2011). Therefore, superiority of FEST has previously been shown in larger studies, while cortical mechanism remained unclear. Our study utilized detailed assessments with an individual suffering mild upper-limb motor impairment after chronic TBI to understand mechanisms of recovery and time course of cortical changes after FEST. As recently pointed out, case study observations utilizing detailed aspects of intervention can serve as a basis for future studies targeting larger populations (Bloem et al. 2020). Therefore, our current study results should be used to develop specific hypotheses for the future studies related to cortical mechanisms of motor improvement using FEST after TBI.

## 5. Conclusion

Our clinical case study results showed that an upper-limb FEST intervention can be effective for eliciting cortical re-organization that can improve voluntary upper-limb fine motor function of an individual suffering from mild motor impairment resulting from chronic TBI. Our study showed that motor changes were related to cortical re-organization, consistent to previously shown clinical carry-over effects (Kapadia et al. 2011). Specifically, we showed that 12-weeks of FEST, which included 36 sessions lasting 45-60 min of task-specific and repetitive FES-assisted reaching and grasping, can elicit long-term facilitation of corticospinal excitability, likely due to larger motor map representations in and around the primary motor cortex. Increased activations after FEST were also shown in the somatosensory areas, as well as other areas related to voluntary motor control and sensorimotor integration, suggesting widespread cortical re-organization. Assessments also suggested that cortical changes may persist after the intervention. The mechanism of long-term FEST elicited cortical re-organization likely involve integration of voluntary motor commands and sensorimotor network engagement through FES. Overall, our study showed evidence that FEST can be applied in chronic stage TBI to elicit cortical re-organization and improve fine motor function.

## Data Availability

The data that support the findings of this study are available from the corresponding author, upon reasonable request.

## Acknowledgments

The authors would like to thank Mr. Daiju Ikawa and Mr. Yutaka Tazawa for their help during the study.

## Funding

The project was funded by the Japan Society for the Promotion of Science Grants-in-Aid for Scientific Research: KAKENHI (Grant #: 18H04082, 18KK0272, 19K23606, and 20K19412).

## Conflict of interest

M.R.P. is a shareholder and director in company MyndTec Inc. The remaining authors have no conflicts of interest.

## Notes

### Competing Interest Statement

M.R.P. is a shareholder in company MyndTec Inc. The remaining authors have no conflicts of interest.

### Funding Statement

This project was funded by the Japan Society for the Promotion of Science Grants-in-Aid for Scientific Research - KAKENHI (Grant numbers: 18H04082, 18KK0272, 19K23606, and 20K19412).

### Author Declarations

The study was approved by the local institutional research ethics committee at the University of Tokyo.

### Summary of Updates

Statistical analysis results added.

