## Supplementary information for "Cortical re-organization after traumatic brain injury elicited using functional electrical stimulation therapy: A case report"

Milosevic et al.

**Supplementary information**

***Patient history***

In the initial assessment, which was administered after the accident, the participant was diagnosed as having suffered a diffuse brain injury, multiple trauma, skull fracture, pulmonary contusion, and hemorrhagic shock. At the time of injury, the participant was diagnosed as a serious condition by the Glasgow Coma Scale (GCS) (Teasdale et al. 1974): no eye opening, no verbal response, and no motor response. Physiological testing concluded that there was no injury to the spinal cord. The participant received skull reconstructive surgery and remained in intensive care unit for 3-weeks, which was followed by one-month of monitoring, before 5-months of inpatient rehabilitation, where he received standard rehabilitation for 3-hours per day. After discharge, he was still unable to walk independently and required assistance during daily living. Over the ensuing six years, he continued various rehabilitation and training programs, including Pilates and brain gymnastics. Ultimately his lower-limb function improved, and he was able to walk independently, while his upper-limb impairment persisted.

At the study onset, symptoms related to movement function included: (1) ataxia, specifically characterized by tremor in the right upper- and lower-limbs (i.e., contralateral to the trauma) during movement initiation, as well as trunk, whole body movement, and balance disorders; (2) involuntary movements in the right thumb, and tremor during performance of fine motor tasks such as writing and using chopsticks; (3) mild hemiplegia mainly affected the right foot; and (4) eye movement disorder, characterized by poor eye movement control. Symptoms related to higher brain dysfunction, included: (1) memory loss, related to pre-accident and recall of new events after the accident; (2) attention disorder, characterized by decline in arousal, decline in attention (sleeping or drowsiness), specifically during multi-tasking activities; (3) performance impairment, including impulsive behaviour; and (4) social behavior disorders, characterized by decline in recognition of anger and emotional control.

***FES***

Each training session consisted of three functional training protocols, as illustrated in Figure 1B: (1) palmar grasp - to generate hand opening, a cathode was placed on the wrist extensors (extensor carpi radialis: 19.6±4.2 mA) and the anode on the extensor tendons (dorsal side of the wrist); to generate a palmar grasp, the cathodes were placed on the thumb (abductor pollicis brevis: 9.6±2.4 mA) and wrist flexors (flexor carpus radialis: 9.6±2.9 mA and flexor carpus ulnaris: 10.5±3.1 mA) and the anodes on the flexor tendons (palmar side of the wrist); (2) hand-mouth - to generate elbow and shoulder flexion, the cathodes were placed on the biceps (biceps brachii: 17.8±6.1 mA) and shoulder (anterior deltoid: 15.7±4.5 mA) with the anode also placed on the muscle belly away from the cathodes; to generate elbow and shoulder extension, the cathodes were placed on the triceps (triceps brachii: 18.8±4.5 mA) and the shoulder (posterior deltoid: 21.3±4.7 mA) and the anodes on the muscle belly away from the cathodes; and (3) point forward - to generate hand pointing forward, the cathodes were placed on the triceps (triceps brachii: 18.2±2.7 mA) and shoulder (anterior deltoid: 17.1±3.9 mA) with the anodes on the muscle belly away from the cathodes; to generate hand retraction, the cathodes were placed on the biceps (biceps brachii: 16.9±5.2 mA) and shoulder (posterior deltoid: 21.2±4.6 mA) and the anodes on the muscle belly away from the cathodes (Figure 1B). Each movement was delivered independently during the sessions. In each protocol, participant performed a specific functional task, including grasping a water bottle (palmar grasp), bringing an object to their mouth (hand-mouth), and pointing towards a target (pointing forward). For each trial, the participant was first asked to attempt to perform the task himself without the help if FES, while the therapist triggered a pre-programmed FES sequence after allowing the participant to initiate the movements to assist his voluntary effort.

***fMRI data acquisition***

During fMRI assessments, the target force trajectories consisted of four phases: rest (10 sec), ascending (10 sec), keep at 20% MVC (10 sec), and descending (10 sec) (Kuhtz-Buschbeck et al. 2001). fMRI scan sessions were repeated four times for each task and averaged for the four hand grip and four finger pinch tasks. A rest period of at least 20 sec was given between each trial. Force data was recorded using a custom program written in LabVIEW (National Instruments, Austin, TX, USA) and digitized at 1,000 Hz sampling frequency using an analog-to-digital converter (USB-6259 BNC, National Instruments, Austin, TX, USA). Force data during fMRI sessions was used to ensure that the participant was following the target force trajectories during fMRI scans.

All MRI images were acquired using a 3T MRI scanner with a 64-channel head coil (MAGNETOM Plisma, Siemens, Germany). Functional T2*-weighted echo-planar images to reflect blood oxygenation level-dependent (BOLD) responses (Ogawa et al. 1990) were collected using the following parameters: TR=2,000 ms, TE=25 ms, flip angle=90°, FOV=192 mm, 39 contiguous axial slices acquired in interleaved order, thickness=3.0 mm, in-plane resolution = 3.0×3.0 mm, bandwidth =1,776 Hz/pixel, as in previous studies using similar force match tasks (Noble et al. 2013; Naito and Hirose 2014). Auto-align was run at the start of each session. High-resolution T1-weighted structural images were also acquired, using the 3D MPRAGE (T1-weighted anatomical images) pulse sequence: TR=2,000 ms, TE=2.9 ms, flip angle=9.0°, FOV=256 mm, 176 contiguous axial slices, thickness = 1.0 mm, in-plane resolution: 1.0×1.0 mm (Noble et al. 2013; Naito and Hirose 2014).

***fMRI data processing***

Prior to data analysis, DICOM image files were converted to NIFTI format. First, preprocessing was performed in the following order: (1) Realignment - excessive head movement was corrected using the realignment procedure by applying a threshold of 2 mm for translation and 2° for rotation (NOTE: since no excessive movements were identified in any of the images, no scans were excluded); (2) Coregistration - the T1-weighted structural scan and the average EPI-scan in each of the four experimental conditions were aligned to superimpose the head position information; (3) Normalization - segmentation of the structural scan was performed, providing normalization parameters, which were used to normalize the EPI-scans to the Montreal Neurological Institute (MNI) space (resized voxels 3×3×3 mm) (Kuhtz-Buschbeck et al. 2001); (4) Smoothing - EPI-scans were smoothed with a Gaussian kernel of 8 mm; and (5) Scaling - the value in each voxel was normalized by converting it into a percent signal change (PSC), which was the percentage increase from the mean of the whole brain in each session and an indicator of the intensity of the BOLD signal (Noble et al. 2013). The PSC value was calculated on a voxel-wise basis, for each condition.

***Supplementary references***

Kuhtz-Buschbeck, J. P., Ehrsson, H. H., & Forssberg, H. (2001). Human brain activity in the control of fine static precision grip forces: an fMRI study. *Eur J Neurosci, 14*(2), 382-390. doi: 10.1046/j.0953-816x.2001.01639.x

Noble, J. W., Eng, J. J., & Boyd, L. A. (2013). Effect of Visual Feedback on Brain Activation During Motor Tasks: An fMRI Study. *Motor Control, 17*(3), 298-312. doi: 10.1123/Mcj.17.3.298

Ogawa, S., Lee, T. M., Nayak, A. S., & Glynn, P. (1990). Oxygenation-Sensitive Contrast in Magnetic-Resonance Image of Rodent Brain at High Magnetic-Fields. *Magnetic Resonance in Medicine, 14*(1), 68-78. doi: DOI 10.1002/mrm.1910140108

Teasdale, G., & Jennett, B. (1974). Assessment of coma and impaired consciousness. A practical scale. *Lancet, 2*(7872), 81-84. doi: 10.1016/s0140-6736(74)91639-0
